# In Gossip, We Trust: Residents’ Understanding of Gossip as a Social Resource

**DOI:** 10.1101/2025.06.06.25329150

**Authors:** Laura Chiel, Michael Fishman, Lorelei Lingard, Erik Driessen, Emmaline Brouwer

## Abstract

**Introduction:** Gossip is pervasive in residency programs and may play a key role in resident development. Recommendations for how to deal with harmful gossip in residency programs have been proposed but, before addressing gossip, we need to more fully understand how gossip in residency programs functions as a social process and explore residents’ experiences with gossip across contexts. In this study, conducted from a constructivist vantage point, we aim to explore residents’ experience of gossip in the residency workplace.

**Methods:** Constructivist grounded theory was used to iteratively conduct and analyze interviews with 16 resident participants from pediatric, internal medicine, obstetrics-gynecology, and psychiatry programs located in the United States and the Netherlands. Interview questions focused on residents’ personal experiences with gossip in training.

**Results:** We found that gossip has multiple emotional impacts on participants, while also helping them navigate the learning environment. Gossip participation itself must be navigated, but how participants do so varies, with each following a different map, or unspoken rules surrounding gossip etiquette, often routed by social connectivity and trust.

**Discussion:** We conceptualize gossip as a social resource in residency training. Gossip influences residents’ emotions and, through gossip, residents learn what is expected of them and from others. However, gossip is not uniformly available. Residents likely experience differential emotional support and requisite information gained through gossip. Because such support and information are critical in residency, program leaders should be aware of the influence of gossip and seek to understand social connectivity in their programs.

## Introduction

Gossip is pervasive in residency programs but until recently we could say little about it and do even less. This is because gossip occurs ‘behind closed doors’ and in the ‘unmanaged spaces’ of organizations’” [1–4]. Gossip in residency programs has recently become a focus of inquiry, and we are beginning to realize its impact on learners [5–9]. What is not yet well understood is how gossip in residency programs functions as a social process – in particular, how the resident experience is influenced by the gossip that diffuses through it.

Gossip in residency training programs, thus far, has been approached as something that can be broken into its component parts which can then be solved. For instance, Chaikof and colleagues focus on the specific ways in which the professional hierarchy interfaces with residents’ experience of gossip [6]. L’Huillier and colleagues create a linear process model of gossip in residency and call to reduce harmful gossip in training programs, proposing strategies such as: ‘commit to transparency when possible’ and ‘encourage mindfulness and emotional intelligence’ [7]. These studies begin to unpack and problematize gossip in residency and move programs towards next steps but mau underestimate its complexity, risking yielding insufficient or even problematic solutions [10].

By contrast, constructivist perspective [11] approaches gossip in residency training as a complex, socially-constructed process that is messy, unpredictable and varies by context. In this qualitative study using constructivist grounded theory (CGT) [12], we approach gossip from this orientation, interviewing residents across settings and specialties to explore, ‘What is residents’ experience of gossip in the residency training environment?’

## Methods

### Study design

We approached the research question from a constructivist vantage point [11], recognizing that individuals make meaning from gossip encounters. We selected CGT methodology [12] to delve into the complex social process of gossip in the residency workplace. In keeping with Charmaz’s conceptualization of CGT [13–15], we seek to interpret and highlight the voices of the study participants themselves.

We provided participants with a definition of gossip, recognizing that gossip would likely be defined differently by each participant. We reviewed the literature for definitions and selected Eder and Enke’s definition of gossip as ‘evaluative talk about a person who is not present,’ given that this definition is broad – assuming no specific intent of the gossiper and without guardrails on the valence of the content – but is somewhat constrained – limited to talk, not writing, and specifically *evaluative* talk about others – and given that we expected this definition to resonate with participants in the study [16–18].

This study is part of a larger program of research on gossip in residency training that includes exploring the interplay between feedback in gossip in residency training.

### Data collection

We used semi-structured interviews to match with the personal nature of the subject. We designed the interview guide to elicit rich experiences with gossip in residency training, including by asking for examples of gossip in the residency training program, times when participants engaged in gossip, and instances when participants have been impacted by gossip during residency. We piloted the guide with two recent residency graduates and updated the questions for clarity (Supplemental Material).

Interviews were conducted by 4 members of the research team between November 2024 and April 2025. Interviewers did not interview participants in their training program to allow for ease of discussion of a potentially sensitive topic. Interviews were conducted on Zoom and recorded and transcribed via Amazon Transcribe. Interview duration ranged from 34-54 minutes, excluding study introduction. Data collection was complete when the study team noted that identified concepts were already well reflected in the final interviews.

### Participants

We initially recruited participants from 5 pediatric residency programs of medium (between 31-60 residents) and large (>60 residents) sizes [19] and geographies (from 5 different states in the United States (US), spanning across the country), seeking both a broad perspective on gossip in residency training programs, while exploring gossip in a field where it is underexplored.

Residency program directors forwarded a study recruitment email to all residents in their programs. We initially purposively sampled to include participants from all residency training years and to represent the involved programs. Residents from all 5 programs responded to the initial request for participation with interest; ultimately only residents from 3 programs returned requests to schedule interviews. Ten pediatrics residents from 3 programs were interviewed.

Based on iterative analysis of transcripts, we next theoretically sampled for residents from other specialties and geographies and for additional male residents to better contextualize transferability of our findings. We introduced new questions into the interview guide focused on permissibility of gossip with various members of the team. We interviewed 3 internal medicine residents, 1 obstetrics-gynecology resident, 1 psychiatry resident, and 1 additional pediatrics resident, for a total of 16 interviews.

In all, we interviewed 4 first-year residents, 4 second-year residents, 7 third-year residents, and 1 fourth-year resident. Nine of the participants identify as women, 7 as men. Fourteen of the participants are training in the US and 2 from the Netherlands. We agreed to not reveal program names given the sensitive nature of the topic and the ethical requirement to safeguard anonymity. Participants were offered a small gift card as a token of gratitude for their contributions.

### Data analysis

LC coded the initial two interviews and shared an initial codebook with the study team, who provided input based on review of these transcripts. Further interview transcripts were then iteratively analyzed and coded by LC in ATLAS.ti, performing constant comparison, and repetitively analyzing earlier transcripts to see how codes’ meaning evolved. All study team members read several additional transcripts to further familiarize themselves with the data during this time. As concepts were developed, the team returned to the codes to understand which codes were and were not represented and further refined the concepts. We iteratively developed the interview guide to deepen our understanding. The study team met online and corresponded by email frequently to distill the concepts and, together, to conceptualize gossip in residency training based on the analysis.

The author team practiced reflexivity, bringing their personal relationships with gossip to the analysis and noting how their own stances influenced interpretation of the findings. LC is the granddaughter of a Rabbi and that gossip is sinful was ingrained in her from an early age [20]; however, during residency she found gossip to be therapeutic and engaged in it readily, albeit walking away from gossip conversations with guilt. Now, as a residency Associate Program Director, she recognizes that residents engage in gossip but does not understand their relationship to it. MF is a trainee and finds gossip to be helpful in preparation for working with others. LL,

ED, and EB have not had the experience of gossiping in residency but are aware of its potential effects in their areas of academic focus, including teamwork, communication, and workplace learning and assessment.

The research qualified for Institutional Review Board exemption at Boston Children’s Hospital (IRB-P00049211).

## Results

We found that gossip has multiple emotional impacts on participants, while also helping them navigate the learning environment. Gossip participation itself must be navigated, but how participants do so varies, with each following a different map, or unspoken rules surrounding gossip etiquette, often routed by social connectivity and trust.

### Gossip has multiple emotional impacts on residents

Gossip influences residents’ emotions, including through supporting bonding, allowing space for venting, and securing validation, while also leading to discomfort, guilt, and self-doubt. Gossip was described as a result of residents’ ‘*trauma bonding or time bonding’* (P1). Gossip can be common between residents due ‘*to feeling like the residents are the people who like really understand your experience’* (P5). A participant explained, ‘*You become really close friends with these people and that professionalism component of it all gets a little blurry [*…*] and so maybe there’s greater comfort in the gossip and a little bit lower of a barrier of professionalism*’ (P1).

While often shared between friends, gossip itself supports new friendships, with a participant explaining, ‘*bonding requires that you put something in and someone else put something in and you’re able to, you know, join together*,*’* elaborating, ‘*And I’m kind of grateful for gossip being a way that like, you’re able to sort of like kind of move past just being your coresident on a rotation and into something that’s more like friends’ (P4*). Whether as an activity between friends or one that builds friendship, gossip helps residents feel socially secure.

In addition to supporting bonding, participants describe gossip as *‘therapeutic’* (P10), a *‘pressure release’* (P1), ‘*cathartic’* (P6), and as a means to secure validation from others. A participant described, ‘*I think it’s very helpful to [*…*] relieve my stress about something, to be able to talk to someone who understands who have gone through a similar path as me’* (P10). Gossip can support *‘validation’* and *‘commiseration’*, with a participant elaborating, ‘*In the ER [Emergency Room] I had an attending who just, I felt like, was very harsh, very blunt and she walked away and I, like, looked to two of the nurses and I was like, is it me? And they were like, no, they were like none of us like her. That’s just how she is*.*’* (P2). Another participant described that this validation can be done ‘*in the most unhealthy ways*,*’* explaining, *‘Oh God, it makes us feel better about ourselves [*…*] I think the residents on the whole have lots of confidence issues and one of the ways they boost their confidence is like making comparisons to others’* (P1). While recognizing beneficial emotional aspects of gossip, this participant points to emotional complexities that gossip may engender.

In contrast to its beneficial impacts, gossip can also lead to discomfort, guilt, and self-doubt. Participants described both general and professional discomfort with gossip. One participant noted, ‘*Just as a human being, we should strive not to gossip as much*,*’* (P9) while others voiced concern about gossiping about patients and families specifically. Engaging in gossip could lead to guilt, with a participant explaining ‘*If I had to go back, I wouldn’t do that again, cause there was no fruit to it, even though it just felt good to do. So yeah [* …*] like a lot of things that probably aren’t good, it felt nice in the moment, but then long term felt like remorse for it’* (P11). One participant described how gossip’s content modulates one’s comfort with it, elaborating, ‘*sometimes it’s really harmless and it can make you feel better. But you know, the gossip that goes really behind someone’s back and with a negative connotation to it, I really, I really try to refrain from that and I really don’t like it and I would also hate if people would do that when I’m not around’* (P12), alluding to participants’ fear about being gossiped about.

Participants described fear of and experience with being gossiped about and gossip leading them to question themselves. A participant described discomfort when an attending gossiped to them about another resident, which left them questioning what the attending would later say about the participant: ‘*I was like the fact that you’re going to start saying negative things about someone else. Like I don’t know what you say about me as soon as I walk away’* (P2). Another elaborated on a time when they learned that they had been described as ‘*overconfident’* through gossip and how because of this they wondered whenever they spoke ‘*is this something that comes off as like, overconfident in some way? Is my voice too loud? Am I too happy?’* (P11). Both gossip in theory and actual gossip about participants in practice led them to have feelings of self-doubt.

### Gossip helps residents navigate the learning environment

While gossip influenced participants’ emotional experiences, it also had many practical implications for resident learning; namely, participants learn through gossip how to approach other members of the training environment, including supervising residents, fellows, and attending physicians (referred to throughout as ‘supervisors’), supervisees, and patients.

Residents gossip about supervisors, often when transitioning to a new rotation, as a means of preparation. Gossip about a supervisor could lead to its recipients’ behavioral change. A participant explained that based on knowledge obtained *‘beforehand*,*’* ‘*you can adapt a bit like*,

*OK, I don’t normally do it this strict but because the supervisor I have for this week is usually really strict on this, I can pay a bit more extra attention to this to prevent getting into an awkward or annoying situation’* (P15). While it can be ‘*hard to know if the information you’re receiving is actually accurate, or if it’s the person [gossiper] didn’t like this person and that’s why they’re telling you, or the person like misperceived something the attending said*,*’* one participant described these *‘expectations’* as so helpful that they *‘can’t stop myself from asking’* co-residents for them (P3). Thus, participants valued gossip as guidance, and, in this case, even if imperfect or potentially incorrect.

Gossip could inform participants’ behavior when supervising more junior residents. A participant recounted how they and a co-supervisor learned, through gossip, about the respective interns, or first year doctors in a residency training program, they would be working with, the former with a reputation of being ‘*fantastic*,*’* the latter who ‘*forgets tasks and, like, gets overwhelmed’*; while the participant did not change their practice given they were paired with a strong intern, they recall the co-supervisor planning *‘to make a checklist for every single item that you’re [the intern] doing and I’m gonna make sure it gets done because I’m worried it won’t otherwise’* (P8). Gossip then may lead to changes in supervision and learning opportunities.

Participants’ change in approach secondary to gossip can extend to patient care. A participant explained that if they come on ‘*for a night shift and someone, you know, tells me these stories about a certain patient – I might be much more reluctant than I otherwise normally would to really engage with them over a single nightshift*’ (P5). Participants qualified that information gained from gossip does not interfere with them engaging in an urgent or medically necessary situation, with one participant elaborating ‘*It doesn’t really matter how you feel about the [patient’s] mom. If their child’s in status [epilepticus], you’re gonna be in that room trying to fix that no matter what. But if you need to have a conversation just about [pain medicine] for their comfort, maybe you’re gonna just try to get them the [pain medicine] without going into the room to talk about it’* (P8). Participants expressed concern for *‘bias’* (P9) from these types of conversations and note that when giving sign out they might *‘sugarcoat’* their message to give others insight into challenging social dynamics with patients and caregivers but *‘not be as forthcoming’* with all of the details (P16). While residents rely on gossip as recipients, they also find themselves softening their sentiments when in the role of gossip provider. Participants provide and use gossip in patient care but, again, recognize that gossip is imperfect.

### Gossip must be navigated, but residents follow different maps, or unspoken rules surrounding gossip etiquette, often routed by social connectivity and trust

Recognizing that gossip has beneficial and negative aspects, gossip conversations are carefully regulated through unspoken rules of gossip etiquette which, given their unspoken nature, vary between residents. While these rules varied from participant to participant, one consistent rule was that participants had few qualms about gossiping about attending physicians. Gossiping about an attending may feel cathartic as compared to gossiping about nurses, respiratory therapists, or interns, with a participant explaining, ‘*I think that’s different in some ways than how I feel about when we’re talking about interns because I’m the [senior training year] and he’s the attending. So there’s a different power dynamic and I think I feel more cathartic and less like s***ty’* (P1). While gossiping about attendings is common, the permissibility of gossiping with residents about attendings does not extend to medical students. A participant elaborated, ‘*And sometimes I think that makes the hierarchy really guide who should participate. Because you know, if we’re gonna write an evaluation on our med student and the med student has been like, talking about how they don’t like our attending or something, it doesn’t really look great’* (P4). Violating gossip etiquette could impact a trainee’s reputation, and gossip experience would seem intertwined with hierarchy.

In seeking to distill the interplay between gossip and hierarchy, we found that how residents consider hierarchy in their gossip experiences is not uniform. For example, one hierarchical approach to gossip was described, as follows: ‘*If it’s like an intern talking about an intern, there’s more engagement. But if it’s a senior talking to an intern, there’s less. But when the seniors are complaining about other seniors, they all kind of like go in the circle of like about this person or talking about other individuals’* (P3). This specific understanding of how gossip and hierarchy relate was not shared by all participants. In particular, some participants took issue with gossiping about peers. A participant noted, ‘*I think anytime you’re sort of thrown into something that feels like a bit like a meat grinder, you kind of want to latch on to your allies. So I just feel like that really limits the amount of cross-resident snark that exists, or like talking poorly about people’* (P5). Beyond peers, the acceptability of gossiping about supervisees and nurses varied between participants, while gossip from someone more senior to more junior often led to discomfort.

For some, more than hierarchy, social connectivity and trust dictate gossip appropriateness. Building on the aforementioned concerns about gossiping about peers, a participant explained that peers are not just who you work with by virtue of station but instead, ‘*the other residents in my department are the closest colleagues I have*,*’* elaborating, *‘I really consider it a bit unsavory to gossip about them’* (P12). Closeness, rather than qualifications alone, mediate gossip behavior. A participant shared that during intern year, they and their friends became close with senior residents who they could gossip freely with noting, ‘*I’d say that they would be like a special group in a different class that felt like a similar relation to the people within my class’* (P8). While the hierarchy *‘does kind of set an initial tone*,’ a participant described that other factors, like friendship, can contribute to gossip acceptability, describing a medical student who was friends with a resident from medical school *‘and so it was kind of a different dynamic because they had a social relationship outside of work’* (P4). Trust was repeatedly cited as a key motivator of gossip decisions. A participant explained *‘if you like the coffee lady and you trust her, you can gossip with her*,*’* elaborating, ‘*usually gossip maybe has a bit of negative connotation to it, and I think it doesn’t have to be negative, but when you engage in gossip, you should do it with something that you really confide in, someone who can keep a secret’* (P12). Finally, participants who tend to share their experiences directly with program leadership ‘*tend to not have like the same relationships’* with other residents, lacking someone ‘*you can vent to, and you do that for five minutes, and you might forget about it’* (P8), coming full circle to the affective benefits of gossip.

## Discussion

In this study, we found that gossip influences how residents feel and how residents learn, and the way in which residents access gossip is mediated by hierarchy and unstated rules, often rooted in social connectivity and trust. Our findings, like those of other studies of gossip in residency training, suggest that residents can learn how to participate in the training environment and interface with others through gossip [6, 7]. Our study emphasizes that gossip is not simply a problem to be solved in medical education; it is also a resource through which residents access the hidden curriculum [21]. Our findings further highlight that procuring this resource is complicated, therefore residents devise standards for gossip etiquette that are individualized. In other words, residents must navigate unspoken rules to even access this hidden information. In parallel to seeking to gain entrustment of clinical responsibilities [22], residents must themselves navigate who to trust in obtaining necessary knowledge.

Our findings of the emotional, functional, and ‘hidden’ aspects of gossip have been described in other organizational systems. For example, in the nursing literature gossip has been described as a mechanism to express emotions and as a means of socialization for nurses, albeit one that can lead to guilt [1, 3, 23]. Further, gossip in healthcare organizations has been described as a “reflection of the ‘problem-behind-the-problem,’” allowing leaders who tune in to know “what is ‘really going on’” which, in the case of healthcare organizations, can have meaningful implications for team performance and patient safety [24]. Framed in this way, both how residents access gossip and how program leaders process gossip in their programs have important implications.

We shed light on how residents access gossip and conceptualize gossip as a social resource in residency training programs. Studies on gossip in other disciplines have also described gossip as a ‘social resource,’ one often leveraged based on hierarchical rank [25]. Research on gossip in residency training specifically has identified the intertwined nature of gossip, hierarchy, friendship, and trust, and similarly put a premium on hierarchy as the key determinant of gossip decisions and recommended gossip behavior [6, 7]. Our constructivist approach led us to find social relationships, those more fluid than the rigidity imposed by hierarchy, as at the essence of gossip dynamics. While others have found that gossip leads to friendship, rather than friendship leading to gossip [26], we found that gossip could lead to friendship and friendship could lead to gossip. Thus, in residency training, there are multiple avenues to obtaining the emotional outlet and information conveyed through gossip, but some degree of social savviness is likely required. Residents must understand the local gossip etiquette.

Because equitable access to requisite information and emotional support is critical in residency, programs should be aware of the influence of gossip on individual residents. Programs should seek to work with supervisors to ensure expectations are explicit, in keeping with L’Huillier’s call for transparency [7]. But clear expectations are not enough – expectations will not yield the bonding, commiseration, and other emotional outlets that gossip fulfills. Rather than prioritizing resident-centric strategies focused on burnout and emotional intelligence [7], we recommend strategies that first focus on program leaders themselves, ensuring that they clue into gossip and its underlying dynamics. Program leaders could step back and seek to understand the social networks in a program. Do all residents have someone to turn to? Knowing that negative gossip tends to focus on few individuals, and often those who are least socially connected [27], are there systems to help socially engage residents who may be more likely to be the focus of harmful gossip? Future studies might investigate social networks of gossip in residency training programs.

Limitations of this study include that residents who opted to participate may have been drawn to the study due to a strong viewpoint on gossip. We suspect that this limitation is minimal, given the balanced stances that participants provided, highlighting both beneficial and difficult aspects of gossip. Ours and prior studies have focused on programs in the global west; understanding gossip culture in training programs in other parts of the world would allow for a richer understanding. Further, recognizing that residents’ social connectivity is likely inherently intertwined with their individual identities, the interplay between gossip experience and resident identity is an important avenue for further research.

We found that gossip influences residents’ emotional and educational experiences, but that how to navigate gossip is not clear and that who benefits from the social resource of gossip in residency interplays with their social connectivity and trust. Gossip is quiet -- how to access it, even quieter – leading to opportunity for inequity in our learning spaces.

## Data Availability

All data produced in the present study are available upon reasonable request to the authors

## Acknowledgements

The authors wish to acknowledge Dr. Dorene Balmer for piquing their interest in gossip.

## Supplemental Material

*Let’s start with the day to day – tell me about some examples of gossip in your residency workplace*.

*How did people respond? What did you think of that? What did others’ think?*

*How do you think such instances of gossip are perceived by others in the program? Why do people gossip? Why do you think residents participate in gossip?*

*Tell me about a time you engaged in gossip. How did you come away feeling? What was the outcome of that conversation?*

*Have you been impacted by gossip? Can you tell me more about that?*

*Tell me about a memorable time when gossip was happening in your program. Did the program respond in any way? How did the program respond?*

